# Evaluating the impact of alternative intervention strategies in accelerating onchocerciasis elimination in an area of persistent transmission in the West Region of Cameroon

**DOI:** 10.1101/2022.06.19.22276627

**Authors:** Kareen Atekem, Ruth Dixon, Aude Wilhelm, Benjamin Biholong, Joseph Oye, Hugues Nana Djeunga, Philippe Nwane, Franklin Ayisi, Daniel Boakye, Joseph Kamgno, Elena Schmidt, Rogers Nditanchou, Laura Senyonjo

**Affiliations:** Sightsavers, Cameroon Country Office; Sightsavers, Haywards Heath, United Kingdom; National Programme for the Fight against Onchocerciasis and Lymphatic Filariasis, Ministry of Public Health, Cameroon; Filariasis and other Tropical Diseases Research Center (CRFilMT), Yaoundé, Cameroon; Faculty of Medicine and Biomedical Sciences, University of Yaoundé I, Yaoundé, Cameroon; Parasitology Department, Noguchi Memorial Institute for Medical Research, University of Ghana, Legon, Accra Legon, Accra; The End Fund, New York, USA

**Keywords:** Onchocerciasis, alternative strategies, elimination

## Abstract

**Background:** Alternative strategies are recommended to accelerate onchocerciasis elimination in problematic areas including areas where annual ivermectin (IVM) distributions are unable to interrupt transmission. The aim of this study was to accelerate progress towards elimination in the Massangam health district, West Region of Cameroon where impact evaluations demonstrated ongoing transmission of onchocerciasis infection and high microfilaria (mf) prevalence despite more than 20 years of annual IVM distribution.

**Methodology/Principal findings:** Parasitological, entomological, and breeding site surveys were conducted in 2015 delineating a focus of high transmission and identified three communities with high mf prevalence. Individuals in these communities were screened for mf yearly for a period of two years and those positive treated each year with doxycycline 100mg daily for five weeks. In addition, surrounding communities were given biannual IVM. Temephos-based applications were performed once a week for 10 consecutive weeks on *Simulium* breeding sites. Parasitological and entomological assessments were conducted after two years of implementation and findings compared with 2015 baseline. Alternative strategies accelerated progress towards elimination through a significant mf reduction (χ2: 40.1; p<0.001) from 35.7% (95%CI: 29.0 -42.8) to 12.3% (95%CI: 9.0 - 16.4). Reductions were furthermore recorded over longer time period, with a reduction of mf prevalence by 23.2% following the two years of alternative strategies compared to 20.3% reduction over 15 years of treatment with IVM (1996-2011). Entomological assessment demonstrates that transmission is still ongoing despite the reduction in mf which is expected in an environment with complex breeding sites and open transmission zones.

**Conclusion/Significance:** This study provides evidence that alternative strategies are feasible and effective and should be considered in areas where transmission is sustained throughout long term uninterrupted MDA with IVM. However, there is need to consider wider transmission zones, and further explore optimal timing of larviciding with treatment to impact transmission.

**Author summary:** Elimination of onchocerciasis has showed to be possible when ivermectin (IVM) is given continuously every year for about 15-17 years. However, areas where continuous IVM distribution has not achieved this objective, alternative methods are needed. Massangam health district in Cameroon is one of such areas that IVM treatment has not stopped the spread of the disease despite more than 20 years of annual distribution. This study aimed to fast-track elimination through alternative intervention strategies (AIS). This included testing and treating those having onchocerciasis with doxycycline in communities where the infection was high, giving IVM twice a year to surrounding community members and reducing the flies that carry the worms by pouring chemical in river sites having fly larvae once a week for 10 weeks. The effect of these activities was measured and compared with previous data. The AIS significantly reduced percentage of those having the disease from 35.7% to 12.3%. A 23.2% reduction was also observed with two years of AIS compared with 20.3% reduction with IVM over 15 years. Thus, AIS are practical and useful and should be considered in areas where IVM has not successfully stopped the spread of onchocerciasis.

## Introduction

Onchocerciasis, commonly called river blindness, is a neglected tropical disease (NTD) caused by the parasite *Onchocerca volvulus* (OV). This disease is transmitted through the bite of an infected vector, blackfly of the genus *Simulium,* that breeds in fast-flowing streams and rivers. Adult OV worms produce microfilaria (mf) that move to the skin and eyes causing dermatitis, often with intense pruritis, and if chronic, can lead to skin depigmentation as well as visual impairment including irreversible blindness (1). Currently, about 218 million people live in areas known to be endemic for onchocerciasis, including Cameroon (2).

The current global strategy to eliminate this disease has been the interruption of transmission through mass drug administration (MDA) of ivermectin (IVM) to the eligible populations in endemic areas using the Community-Directed Treatment with Ivermectin (CDTi) approach. It has been demonstrated in a number of countries, including Mali and Senegal, that elimination is achievable after about 15-17 years of annual uninterrupted distribution of IVM alone (3, 4). However, there are other areas where this strategy has not achieved elimination due to low therapeutic coverage (5) or high pre-intervention endemicity and perennial transmission. Further, there are also areas that had been excluded from the interventions or started treatment late, especially hypo-endemic areas (6-8). In addition, areas co-endemic with loiasis, where individuals cannot be treated with IVM due to risk of severe adverse effects (SAEs) (9, 10) pose a significant elimination challenge. A number of programmatic factors, such as scarcity of expert entomologists, difficulties in mapping hypo-endemic areas, and suboptimal diagnostic tests also hinder global progress towards elimination (11).

The World Health Organisation (WHO) roadmap for NTDs sets up a target of increasing the number of countries verified for interruption of transmission of onchocerciasis from 4 (12%) in 2020 to 12 (31%) in 2030 (12). To accelerate the achievement of this ambitious goal, the WHO recommends to use alternative intervention strategies (AIS) in problematic areas, where traditional IVM MDA has shown to be suboptimal (13). These AIS include complementary vector control, biannual or pluriannual MDA, better timing of MDA, and test and treat preventive chemotherapy with alternative drugs (13, 14).

In the West Region of Cameroon, an area of persistent high transmission was identified following an impact evaluation in 2011 that demonstrated a high degree of ongoing transmission after more than 15 years of annual treatment with IVM (15). Operational research was undertaken to determine factors related to failure to achieve elimination thresholds (16). While IVM MDA coverage was generally around the target (above 80% therapeutic coverage), it was reported that about 7.4% of the eligible population were systematically untreated, i.e., had never taken IVM. Entomological and epidemiological surveys in 2015 and 2016 delineated a focus of high transmission within approximately 12km radius with three communities, Makouopsap, Makankoun and Njinja/Njinguoet having mf prevalence of 37.1%, 36.8% and 27.5% in adults aged >15 years and seroprevalence of anti-OV antibodies of 59.0%, 32.4% and 6.2% in children 3-9 years, respectively. This was facilitated by productive perennial blackfly breeding sites on the Nja and Mbam rivers. With these, it was concluded that annual MDA with IVM alone was not sufficient to interrupt transmission (17).

In collaboration with the Ministry of Health (MoH), it was decided that alternative strategies would be required to accelerate progress towards elimination of onchocerciasis in this area and three strategies which included testing individuals for onchocerciasis and treat positive individuals with doxycycline, biannual MDA, and ground larviciding of blackfly breeding sites were considered. In this paper, we describe the implementation and evaluate the impact of these strategies.

## Methods

### Study area

The study was conducted within and around a high transmission area inside Massangam health district (HD) encompassing three focal communities of Makouopsap, Makankoun and Njinja/Njinguoet (Fig 1). The Massangam HD has two main rivers, the Noun which lies to the West and the Mbam to the East, with the latter known to have the most productive breeding sites for *Simulium* vectors (18). This area is characterised by mountainous terrain and constitutes a watershed from which many tributaries of the river Mbam rise, mainly rivers Nja and Kim. The main activity in this rural area is farming for the settled (main/native) communities, with the majority of the farmers moving closer to the rivers during farming seasons thus increasing contact between them and the vector blackflies. In addition to farming, there is cattle rearing practiced primarily by semi-nomadic population subgroups, who tend to live in remote settlements and have mobile lifestyles.

**Fig 1.**
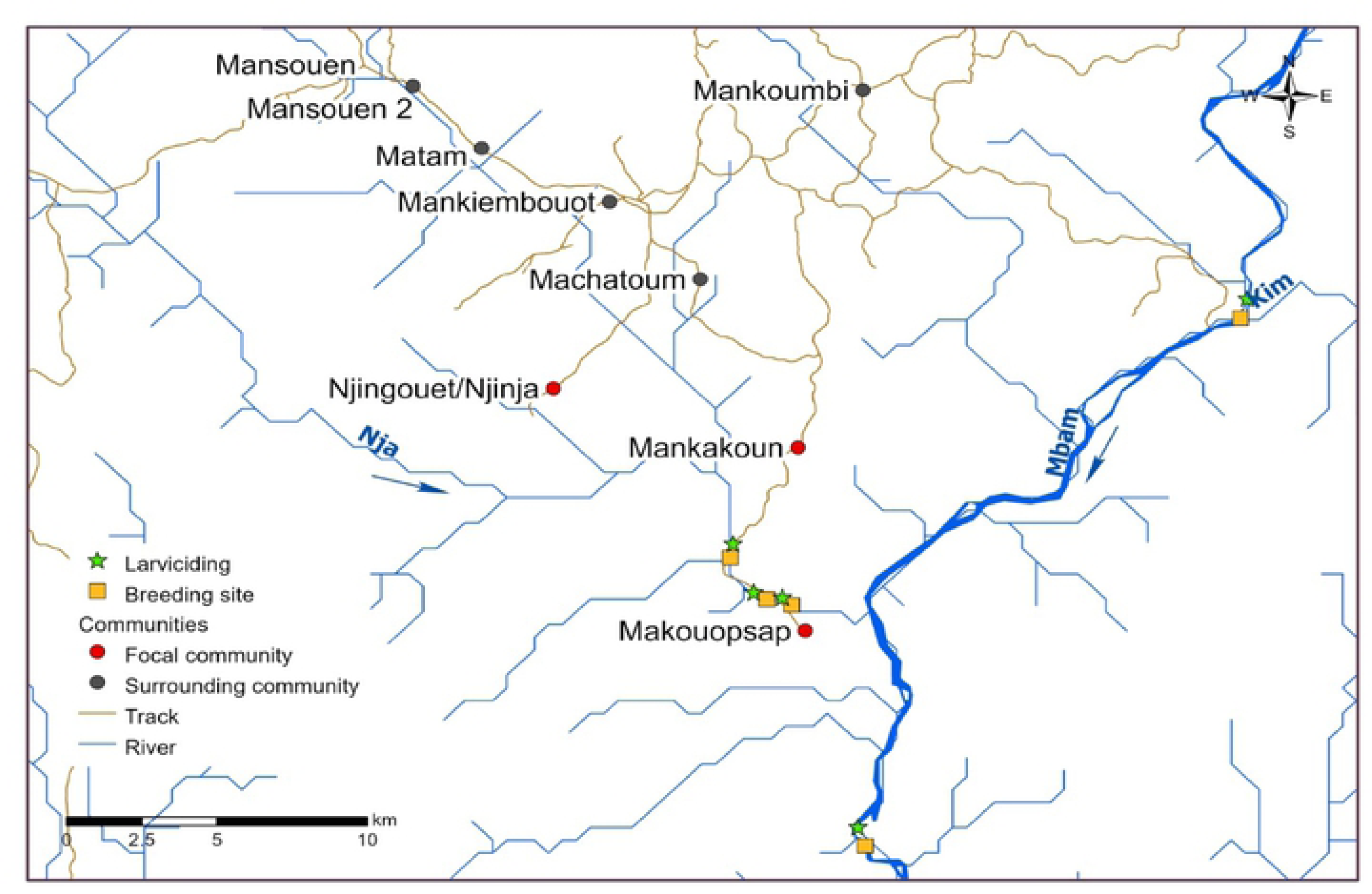
Map showing the focal and surrounding communities, river network, breeding sites and points of larviciding.

The area has had MDAs with IVM since 1996 with reported programmatic coverage of above 80% for five consecutive years prior to the AIS implementation (19).

### Ethics

This study was reviewed and approved by the *Comite National d’éthique de la Recherche pour la Santé Humaine* in Cameroon (approval number 2017/06/918/CE/CNERSH/SP). An administrative authorization was also obtained from the MoH (approval number 631–1817). A written informed consent was obtained from all participants aged 18 and above and a written parental consent and individual assent where appropriate, was obtained from all individuals aged below 18 years.

### Study design

This was an implementation research nested within the National NTDs Programme, which implemented three alternative strategies within and around the identified focus area of high transmission. The outcome was evaluated using surveys of prevalence and intensity of *O. volvulus* infection in humans and the infection and infectivity rates of flies.

### Implementation of the alternative intervention package

The package of strategies was designed and implemented by the National NTDs Programme together with Sightsavers. Each element is outlined below with further details of methods and rationale in table 1.

**Table 1:** Implementation of the various AIS package.

| **Strategy**<br>**(Intervention)** | **Implementation**<br>**site** | **Rationale for intervention** | **Eligibility** | **Timing** | **Method** |
| --- | --- | --- | --- | --- | --- |
| **Doxycycline**<br>**test and treat** | Three focal<br>communities | Doxycycline is a<br>macrofilaricide as compared to<br>IVM which is only a<br>microfilaricide. Two rounds<br>were offered to ensure<br>opportunity for all the<br>population that were initially<br>missed – either due to<br>absenteeism, refusal, or<br>ineligibility | Individuals $\geq 9$ years,<br>not pregnant,<br>breastfeeding or<br>severely ill | Aug 2017 and Sept 2018<br>respecting a minimum<br>duration of 6 months after<br>IVM MDA | All eligible individuals tested for<br>presence of at least one OV mf<br>identified by microscopy in at least one<br>of the two skin snips taken from the<br>iliac crest. If positive, a five-weeks<br>daily dose of doxycycline (100mg) was<br>administered by community drug<br>distributors (CDD) under directly<br>observed therapy (DOT). |
| **Biannual CDTi** | Biannual in seven<br>communities<br>surrounding the<br>focal communities | Perennial transmission in the<br>area, which will not be<br>suppressed by annual IVM<br>alone. Further rounds also help<br>to increase the proportion of<br>individuals who received at<br>least a round of MDA a year | Individuals $\geq 5$ years,<br>not pregnant,<br>breastfeeding for at<br>least 8 days | Sept 2017 (round 1);<br>March 2018 (round 2);<br>Sept 2018 (round 3) and<br>March 2019 (round 4) | Community-directed treatment with<br>ivermectin (CDTi) |
| | Annual in the 3 focal communities | To ensure false negative individuals are also receiving standard CDTi | Individuals $\geq 5$ years, not pregnant, breastfeeding for at least 8 days | Sept 2017 (round 1); March 2019 (round 4) | |
| **Vector control** | Upstream of breeding sites on River Mbam (2) and Nja (3) closest in proximity to focal communities | To reduce fly population and potential for onward transmission of *O. volvulus* | Presence of *Simulium* larvae | Dec 2017 – Jan 2018 and Jan – Feb 2019 (targeting the same breeding sites both rounds and additional breeding site) | Temephos was applied at a dosage of 0.5mg/L (following susceptibility testing) upstream of the identified breeding sites once a week for 10 weeks and fly population monitored weekly and temephos quantities adjusted accordingly. |

### Doxycycline test and treat

Two rounds of test and treat with doxycycline (TTd) were implemented in the three focal communities: with round one in August 2017 (intervention one) and round two in September 2018 (intervention two). This strategy was implemented as doxycycline has been shown to be a macrofilaricide, through the target of the symbiont Wolbachia (20).

Sampling and registration: All individuals within the three focal communities were invited to participate following community mobilisation and sensitization meetings and a community census. Screening teams were set up at centralised points (health centre and residence of community leader) and individuals visited the point closest to their dwelling. For those who did not present at the screening posts, the team members together with Community Drug Distributors (CDDs) did a one-time house-to-house visit to invite them to participate. Upon presentation at the screening point, participants were registered, the survey was described to them, and an informed consent was obtained. The survey tool collected basic demographic data including age, sex, pregnancy status (participant affirmation), CDTi treatment and test and treat participation histories.

Skin snip and microscopy: This was offered to individuals nine years and above, excluding pregnant women. After clinical examination and nodule palpation, the iliac crest skin area was disinfected with cotton dipped in 70 % alcohol. Two skin biopsies, one from each iliac crest were taken using a 2 mm sterile corneo-scleral punch, and the spots were covered with medicated plaster. The skin sample from each participant was placed in two separate wells of a microtitre plate each containing one drop of sterile normal saline. The corresponding well number was reflected on the participant’s identification form. The microtitre plates were sealed with parafilm to prevent any spill over or evaporation and incubated at room temperature for 24 hours. Reading of the slides were conducted in the field. Emerged mf were counted using a light microscope at x10 objective magnification and all mf counts were recorded and expressed per skin snip.

A positive individual, determined as an individual with at least one mf identified in the skin, who was eligible (≥9 years, not pregnant or breastfeeding, and with no underlining chronic health condition), was offered 100mg doxycycline treatment daily for 35 days. The treatment was offered under directly observed therapy (DOT) with food by trained CDDs either through a fixed post or door-to-door distribution. During the treatment course, participants were monitored for any SAEs and compliance to treatment recorded.

The screening and treatment process was repeated the following year. Individuals in these communities (previous and new inhabitants) were (re)screened and any new positives and/or those who remained positive were offered another round of doxycycline treatment as outlined above.

### Ivermectin distribution

Four rounds of CDTi were implemented as biannual IVM rounds every six months for a period of two years as per the WHO guidelines for MDA (21) in the surrounding communities closest to the focal communities – Mansouen 1&2, Mankiembouot, Matam, Machatoum and Mankouombi. It is known that there is perennial transmission of onchocerciasis and one annual round of IVM may not be enough to supress transmission year round.

Those that tested negative for mf or refused to be tested in the three focal communities also received IVM – annually. After conducting census of individuals and households (HH) in these communities, IVM was administered to all eligible individuals, that is, excluding children <5 years, women who were pregnant or those breastfeeding within eight days of delivery, and those severely ill. A door-to-door approach was used, with CDDs using a dose pole to estimate dosage. The HH with absent individuals were visited twice and an individual was recorded as absent if he/she was absent during the second visit. Two coverage evaluation surveys (CES) were conducted to validate treatment coverage achieved for each of the first two rounds of CDTi. Using the sample survey builder, communities were divided into 30 segments of about 50 HH each, and a minimum of seven HH selected per segment. This gave a total of 234 HH and sample size of 1033. Trained surveyors assisted by community guides visited the HH and collected data. Data was recorded electronically using Commcare (https://www.dimagi.com/commcare/), downloaded, cleaned and analysed using the Stata Statistical Software (StataCorp LLC 4905 Lakeway Drive, College Station, Texas 77845 USA http://www.stata.com).

### Vector control

Breeding sites identified in 2015/2016 (17) along the rivers Nja and Mbam and those within and surrounding the focal communities were targeted for larviciding. This strategy was aimed at ensuring the fly population was minimised and timed to after the delivery of the IVM, so as mf in the skin begin to repopulate there is low fly populations for onward transmission. Two rounds of larvicide release (ground larviciding) were carried out in December 2017 to January 2018 and February to April 2019. During the first round, four breeding sites (two on each river) were treated; whilst in the second round, there was an additional breeding site covered on the Nja. These breeding sites were treated once a week for 10 weeks, each year for two years, using temephos at a concentration of 0.5mg/L (determined via susceptibility testing resulting in 100% mortality of blackfly larvae). The impact of larviciding on blackflies population was monitored through two approaches: the regular prospection for larvae at the site of treatment after larviciding and monitoring of adult fly numbers. The latter was through the collection of flies using the human landing collection (HLC) technique at collection points around the riverbanks and communities closer to the breeding sites one day before each weekly larvicide release day. Two trained collectors were positioned per site, and blackflies were caught from 7am to 5pm using locally made sucking tubes (mouth aspirators) as they landed on their exposed legs. The cumulative daily flies were counted and recorded for each catch day and values plotted on a graph to visualize the evolution. Where fly numbers did not decrease in response to larviciding, possible reasons were explored, and adjustments were made to the larviciding strategy as required.

### Impact assessment

#### Parasitological evaluation

A parasitological evaluation was conducted in the three focal communities and one surrounding (sentinel) community, (Mankouombi – which had the highest prevalence among the surrounding communities during the survey in 2015/2016 (17)). This survey was considered as a pre-intervention (baseline) for parasitological comparisons after implementation of the AIS. The impact assessment methodology was therefore conducted in a similar way as baseline – from sampling, through registration, nodule examination and skin snip microscopy.

#### Entomology evaluation

The entomological evaluation was conducted between August and November 2019 (24 days); four months after the final larviciding, five months after the last round of IVM treatment and nine months after the second year of the test and treat implementation to ascertain whether ongoing transmission following implementation of the AIS. Two vector collection sites close to the breeding sites on rivers Mbam and Nja were selected. Additionally, one site per community within Makouopsap and Mankakoun near where both rivers flow were selected based on community information on fly density and bites (Fig 1). Collection of black flies was conducted using the HLC technique, as per previously described. However, the collection period was from 8:00 am to 5:00 pm, with the collectors catching interchangeably from morning to noon and from noon to dusk to minimise collection bias and reduced fatigue.

#### Fly dissection

Flies collected were transported to a field laboratory for dissection. Dissection was done as described by Davies and Crosskey (1992) (22). Briefly, flies were knocked down using chloroform vapour and placed on a glass slide containing normal saline. About 70% of all caught flies were dissected using a dissecting microscope (40x magnification) and checked for parity. Parous flies were further dissected by teasing apart the tissues of the head, thorax, and abdomen sections in search of parasite larvae. Once found, the larval status (L1, L2 or L3), number and location were recorded for calculating the parity, infection, and infective rates.

#### Data analysis

Treatment, parasitological and entomological data were collected on paper, double-entered into Excel and analysed using the STATA statistical package, version 13.0 (TX: StataCorp LP).

For TTd interventions, screening participation rate was calculated as the number of individuals tested by skin snip microscopy divided by the total eligible population registered for the intervention at each stage. Positivity rates were calculated as the proportion of screened population that tested positive. Treatment start and course completion rates were determined by the percentage of eligible infected individuals starting treatment and percentage of those starting treatment that had a full course, respectively.

Reported treatment coverage which was obtained by re-entering the data from CDDs registers to validate the program reported coverage was calculated as the number of individuals treated over the total population according to the CDD census and compared with the program coverage. The percentage of people who received all rounds, or at least two rounds of IVM was calculated. Treatment coverage was calculated as number of individuals treated over the total eligible population and compared with corresponding coverage calculated from the register.

For the impact assessment, since the parasitology pre-intervention study data was a subcomponent of a larger survey, the data was extracted and weighted accordingly (using 2017 community census population estimates). The post-intervention sample community weights were based on the 2018 community census population estimates, both conducted at the time of the surveys.

Three parasitological outcomes were investigated to assess the impact of the intervention – namely nodule prevalence, mf prevalence, and mf infection intensity. Nodule prevalence was calculated as the proportion of examined individuals with one or more nodules. The mf prevalence was calculated as the proportion of skin-snipped individuals who tested mf positive by microscopy. The mf intensity was calculated both as average mf intensity and community microfilaria load (CMFL). Average mf intensity was calculated using the arithmetic mean to include intensity measures of all skin snipped for mf – including those with zero intensity counts - so analysis could be done on original (non-transformed) data. The CMFL was calculated using the geometric means for all individuals aged 20 years and above that were screened for mf, using a log(x+1) transformation to include zero mf intensity counts. The data was strongly positively skewed. As a result, a non-parametric test (two-sample Wilcoxon rank-sum test, or Mann-Whitney U test) was used to compare average mf intensities.

Univariate analyses (chi-squared test or logistic regression) of the before and after intervention samples for nodule prevalence and mf prevalence were conducted, stratifying by age, sex, and location. All significant covariate associations with both primary exposure (i.e., categorization as pre-intervention vs post-intervention participant) and outcome (i.e., nodule or mf prevalence) were controlled for in multivariate logistic regression modelling. For each covariate significantly associated with either the primary exposure or the outcome, logistic regression odds ratios were compared when including and excluding the covariate for a >10% difference in its measure, to inform whether the covariate should also be controlled for in the final logistic regression model.

Entomological indices – namely the monthly biting rate (MBR), parous (parity), infection, infective rates, and the monthly transmission potential (MTP) were estimated. The MBR was calculated as the number of flies caught divided by the number of days of fly collection for that month multiplied by the number of calendar days for the relevant month. The parous rate was calculated as proportion of dissected flies that were considered as parous. The infection rate (%) was calculated as the proportion of parous flies with L1, L2 and/or L3 and infectivity rate as the proportion of the parous flies with L3 in the head (L3H). Since no pool screening was conducted on the caught flies (only dissection to determine the infectivity), the infectivity rate could not be compared with the WHO elimination thresholds.

The MTP was calculated for the respective months using the MBR for the month (all capture sites) multiplied by the number of L3H for that month and divided by the total number of flies dissected.

## Results

### TTd intervention

In intervention round one, 1,921 individuals were registered, of whom 1,211 (63%) were eligible for testing; whilst 2,380 participants were registered in intervention round two, and 1,615 eligible. A total of 643 (53%) and 879 (54%) were screened and OV prevalence of 25% and 29% recorded for rounds one and two respectively, with treatment compliance rates of 93% and 98% (table 2).

**Table 2:** Performance of TTd interventions 1 and 2.

|  | INTERVENTION ONE | INTERVENTION TWO |
| --- | --- | --- |
| **REGISTERED** | 1,921 | 2,380 |
| **ELIGIBLE FOR SCREENING** | 1,211 | 1,615 |
| **TESTED (SKIN SNIP)/SCREENING COVERAGE** | 643 (53%) | 879 (54%) |
| **POSITIVE/ OV PREVALENCE** | 161 (25%) | 251 (29%) |
| **STARTED TREATMENT** | 114 (71%) | 208 (83%) |
| **COMPETED TREATMENT** | 106 (93%) | 203 (98%) |

### IVM MDA

Reported treatment coverages (TC) were lower than the program coverage reported for the corresponding round (table 3). Due to inaccessibility of treatment registers containing data from round one, the reported TC for this round was not calculated. However, program reported TC was 64%. The CES indicated generally lower coverages when compared to both program and register with 55.4% (CI: 44.0-66.3) and 59.0% (CI: 45.5-71.3) in rounds one and two, respectively. The percentage of people who received two rounds of IVM within a year were 59% and 66% for round 2 & 3 and 3 & 4 respectively whilst 82% of people participated in at least one of the three rounds.

**Table 3:** Ivermectin treatment coverages over four treatment rounds.

| CDTi round | Distribution period | Program coverage (%) | Reported (register) coverage (%) | CES (95% CI) |
| --- | --- | --- | --- | --- |
| 1 | September 2017 | 64 | NA | 55.4 (44.0-66.3) |
| 2 | March 2018 | 79 | 71 | 59.0 (45.5-71.3) |
| 3 | September 2018 | 82 | 67 | NA |
| 4 | March 2019 | 81 | 69 | NA |

### Ground larviciding

Round one and two temophos treatments showed a similar pattern of rise and fall of fly population. Immediately after the first week of temephos release, a sharp drop in fly population was observed, which gradually increased as the weeks went by up to week four (Fig 2). The continuous increase in the fly population led to adjustments in the breeding sites targeted and quantity of temephos released, which led to a drop in fly numbers from week four to the end of larviciding.

**Fig 2.**
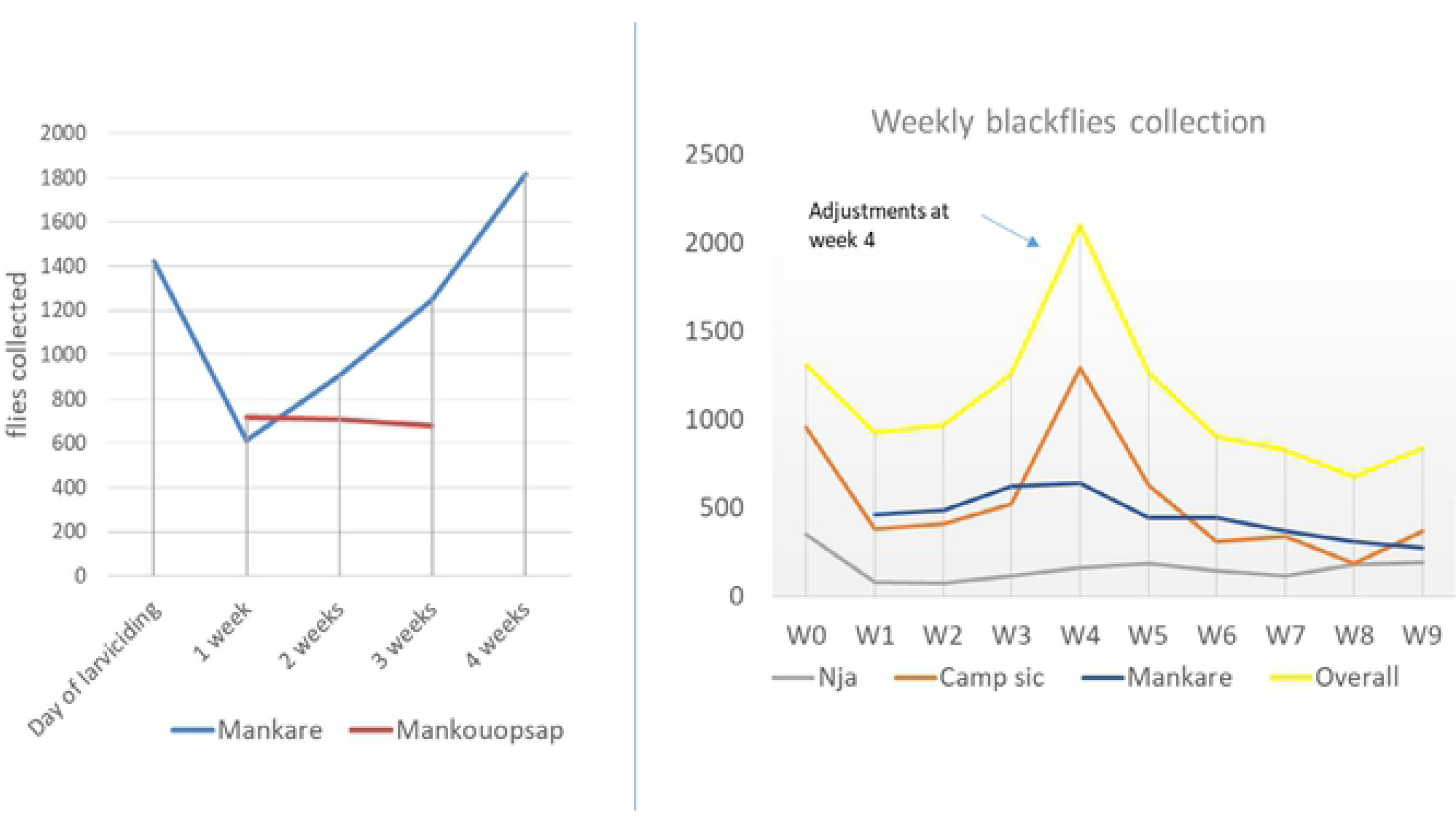
Fly collection during rounds one and two ground larviciding.

### Impact findings

#### Parasitology

##### Nodule Prevalence

A total of 557 and 454 individuals from the three focal communities were enrolled during the pre- and post-interventions respectively and all examined for nodules. Over half of the participants were male (58.2% and 57.7%, respectively). The pre-intervention population was younger with a mean age of 15 years (range 3-90 years) compared to 25 years (range of 3-80 years) during the post-intervention survey. In the sentinel community of Mankouombi, 365 individuals were enrolled and examined during the pre-intervention survey and 292 individuals after the intervention: 52.3% and 57.2% were male. The pre-intervention sample was also younger with the mean age of 19 years (range 3 – 89 years) compared to 39 years (range 5 – 91 years) in the post-intervention period. (table 4).

**Table 4:**
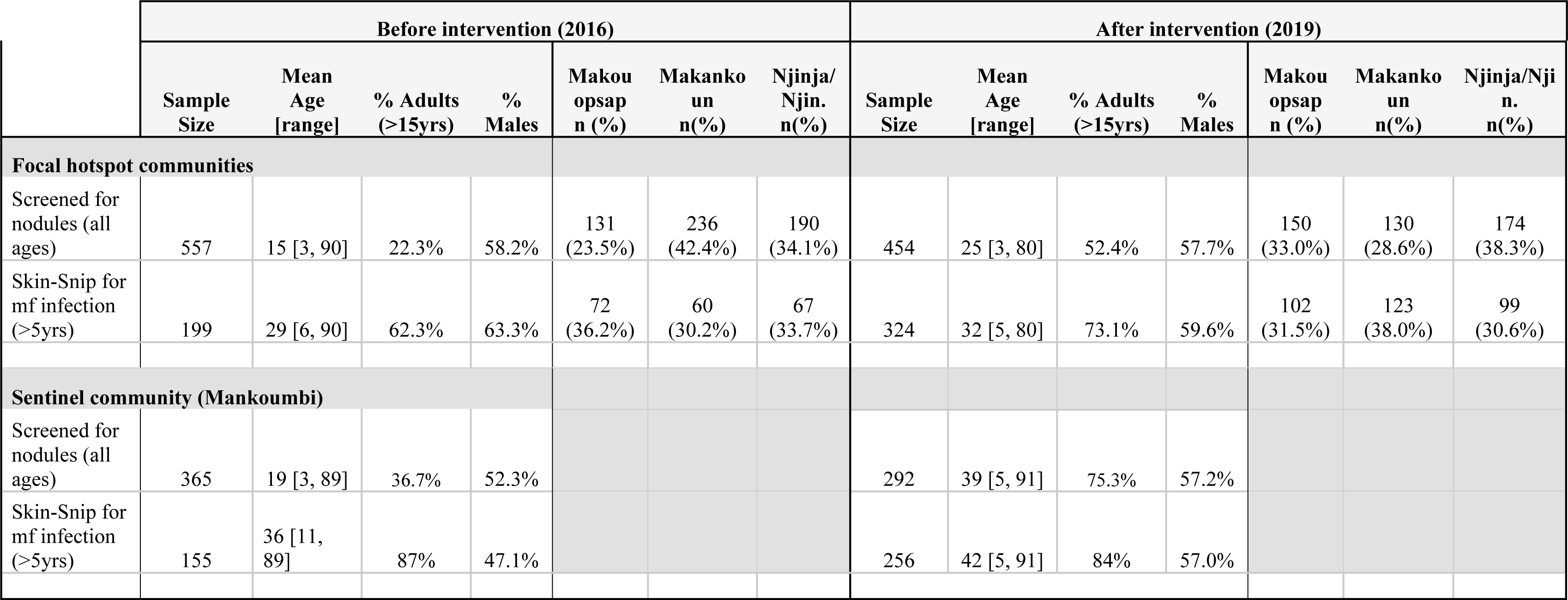
Population description in focal communities.

A multiple logistic regression model controlling for age and community (table 5) found evidence of a difference in overall nodule prevalence after the intervention compared to before the intervention, with an adjusted odds ratio of 0.17 (95% CI: 0.05, 0.54; p=0.01)). There was no significant difference in nodule prevalence between focal communities (Makouopsap versus Njingouet OR=3.40, p=0.11; Mankakoun versus Njingouet OR=1.93, p=0.31), though there was an increased odds of having nodules in adults as compared to children under 15 (OR=7.43, 95% CI: 2.66, 10.75, p<0.01).

**Table 5:**
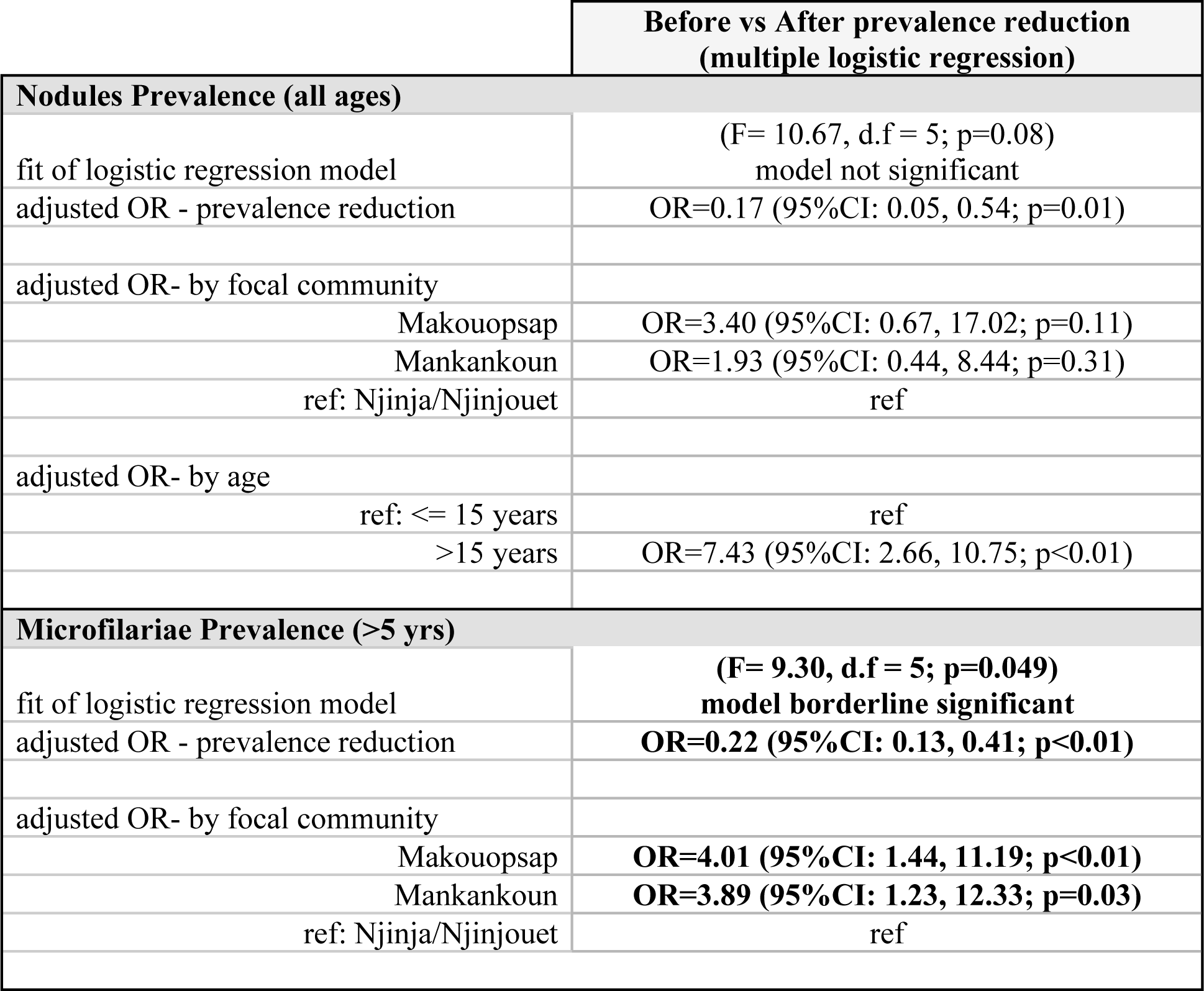
Modelling the significance of prevalence reduction from pre- to post-intervention surveys.

|  |  | **Before vs After prevalence reduction**<br>**(multiple logistic regression)** |
| --- | --- | --- |
| **Nodules Prevalence (all ages)** |  |  |
| fit of logistic regression model | | (F= 10.67, d.f = 5; $p=0.08$)<br>model not significant |
| adjusted OR - prevalence reduction | | OR=0.17 (95%CI: 0.05, 0.54; $p=0.01$) |
| adjusted OR- by focal community |  |  |
| Makouopsap | | OR=3.40 (95%CI: 0.67, 17.02; $p=0.11$) |
| Mankankoun | | OR=1.93 (95%CI: 0.44, 8.44; $p=0.31$) |
| ref: Njinja/Njinjouet |  | ref |
| adjusted OR- by age |  |  |
| ref: $\leq 15$ years | | ref |
| $>15$ years | | OR=7.43 (95%CI: 2.66, 10.75; $p<0.01$) |
| **Microfilariae Prevalence ($>5$ yrs)** |  |  |
| fit of logistic regression model | | (F= 9.30, d.f = 5; $p=0.049$)<br>model borderline significant |
| adjusted OR - prevalence reduction |  | **OR=0.22 (95%CI: 0.13, 0.41; $p<0.01$)** |
| adjusted OR- by focal community |  |  |
| Makouopsap |  | **OR=4.01 (95%CI: 1.44, 11.19; $p<0.01$)** |
| Mankankoun |  | **OR=3.89 (95%CI: 1.23, 12.33; $p=0.03$)** |
| ref: Njinja/Njinjouet |  | ref |

In the sentinel community, which received two rounds of MDA IVM but no test and treat with doxycycline intervention, nodule prevalence before the intervention was 2.5% (95%CI: 1.1 - 4.6), decreasing to 0.7% (95% CI: 0.1 - 2.4), but there is limited evidence of a difference (p=0.07).

##### Microfilaria prevalence and intensity

A total of 199 and 324 individuals (table 6) were skin snipped during the pre- and post-intervention surveys respectively; 63% and 59.6% of which were from men. The mean ages were 29 years (range 6 – 90 years) and 32 years (range 5 – 80 years) respectively.

**Table 6:** Reduction of Nodule and mf prevalence.

|  | **Before intervention (2016)** |  | **After intervention (2019)** |  | **Before vs After** |
| --- | --- | --- | --- | --- | --- |
| **focal communities** | **Total screened** | **% positive**<br>**(95% CI)** | **Total screened** | **% positive**<br>**(95% CI)** | **prevalence reduction**<br>**significance (pearson $\chi^2$)** |
| **Nodule Prevalence - all ages** |  |  |  |  |  |
| Sample Screened for nodules | N = 557 | 9.3% (7.1, 12.1) | N = 454 | 3.7% (2.2, 5.9) | $\chi^2=12.3$ (p<0.001) |
| focal communities |  |  |  |  |  |
| Makouopsap | 131 | 19.1% (12.7, 26.9) | 150 | 3.3% (1.1, 7.6) | $\chi^2=18.2$ (p<0.001) |
| Mankankoun | 236 | 8.5% (5.3, 12.8) | 130 | 3.8% (1.3, 8.7) | $\chi^2=2.8$ (p=0.093) |
| Njinja/Njinjouet | 190 | 3.7% (1.5, 7.4) | 174 | 4.0% (1.6, 8.1) | $\chi^2=0.03$ (p=0.867) |
| **Microfilaria prevalence**<br>**(>5yrs)** |  |  |  |  |  |
| Sample Skin-snipped for mf | n = 199 | 35.7% (29.0, 42.8) | n = 324 | 12.3% (9.0, 16.4) | $\chi^2=40.1$ (p<0.001) |
| focal communities |  |  |  |  |  |
| Makouopsap | 72 | 45.8% (34.0, 58.0) | 102 | 15.7% (9.2, 24.2) | $\chi^2=19.0$ (p<0.001) |
| Mankankoun | 60 | 38.3% (26.1, 51.8) | 123 | 18.7% (12.2, 26.7) | $\chi^2=8.3$ (p=0.004) |
| Njinja/Njinjouet | 67 | 22.4% (13.1, 34.2) | 99 | 1.0% (0.2, 5.5) | $\chi^2=21.0$ (p<0.001) |
| **Sentinel community -**<br>**Mankoumbi** | **Total screened** | **% positive**<br>**(95% CI)** | **Total screened** | **% positive**<br>**(95% CI)** | **prevalence reduction**<br>**significance (fisher's $\chi^2$)** |
| Nodule prevalence | 365 | 2.5% (1.1, 4.6) | 292 | 0.7% (0.1, 2.4) | p=0.07* |
| Mf prevalence | 155 | 11.0% (6.5, 17.0) | 256 | 5.1% (2.7, 8.5) | **$\chi^2=4.95$ (p=0.03)** |

\* 1-sided Fisher's exact test used due to low cell count

**Table 7:** Comparing mf intensity across the surveys.

|  | Before intervention (2016) |  |  |  |  | After intervention (2019) |  |  |  |  | Before vs after |
| --- | --- | --- | --- | --- | --- | --- | --- | --- | --- | --- | --- |
|  | Tested for mf | mf+ | intensity range | CMFL* | average intensity (95% CI) * | Tested for mf | mf+ | intensity range | CMFL* | average intensity (95% CI) * | Average intensity reduction (Mann-Whitney) |
| **Micofilarial Intensity (mf/skin snip; 5yrs+)** |  |  |  |  |  |  |  |  |  |  |  |
| **focal communities\*** | 199 | 71 | [0, 81] | 0.66 | 3.1<br>(-0.4, 6.7) | 324 | 40 | [0, 243] | 0.27 | 2.3<br>(-2.1, 6.8) | Z=6.085 (p<0.001) |
| Makouopsap | 72 | 33 | [0, 50] | 0.57 | 3.4<br>(1.4, 5.5) | 102 | 16 | [0, 187] | 0.22 | 3.8<br>(-0.3, 7.7) |  |
| Mankankoun | 60 | 23 | [0, 81] | 0.89 | 4.5<br>(1.0, 7.9) | 123 | 23 | [0, 243] | 0.49 | 4.5<br>(2.5, 8.1) |  |
| Njinja/Njinjouet | 67 | 15 | [0, 26] | 0.56 | 1.5<br>(0.4, 2.7) | 99 | 1 | [0, 25] | 0.06 | 0.3<br>(-0.2, 0.8) |  |
| **Sentinel community** |  |  |  |  |  |  |  |  |  |  |  |
| Mankoumbi | 155 | 17 | [0, 142] | 0.13 | 1.3<br>(-0.5, 3.1) | 256 | 13 | [0, 58] | 0.11 | 0.7<br>(0.1, 1.4) | Z=2.224 (p=0.026) |

*\*average mf intensity is arithmetic mean of all screened for mf, including zero intensity counts; CMFL is geometric means for ages 20yrs+ and includes all zero counts through use of log(x+1) transformation*

In the sentinel community, 155 and 256 individuals were skin snipped during the pre- and post-interventions; 47.1% and 57% were male and mean ages were 36 years (range 11 – 89 years) and 42 years (range 5 – 91 years).

Dropout rates between population enrolled and screened before and after interventions were calculated (Fig 3). For both focal and sentinel communities, a dropout rate of more than 50% was observed pre-intervention from enrolment to testing, while lower percentage dropouts were recorded during the post-intervention (28.6% and 12.3% for focal and sentinel communities respectively).

**Fig 3:**
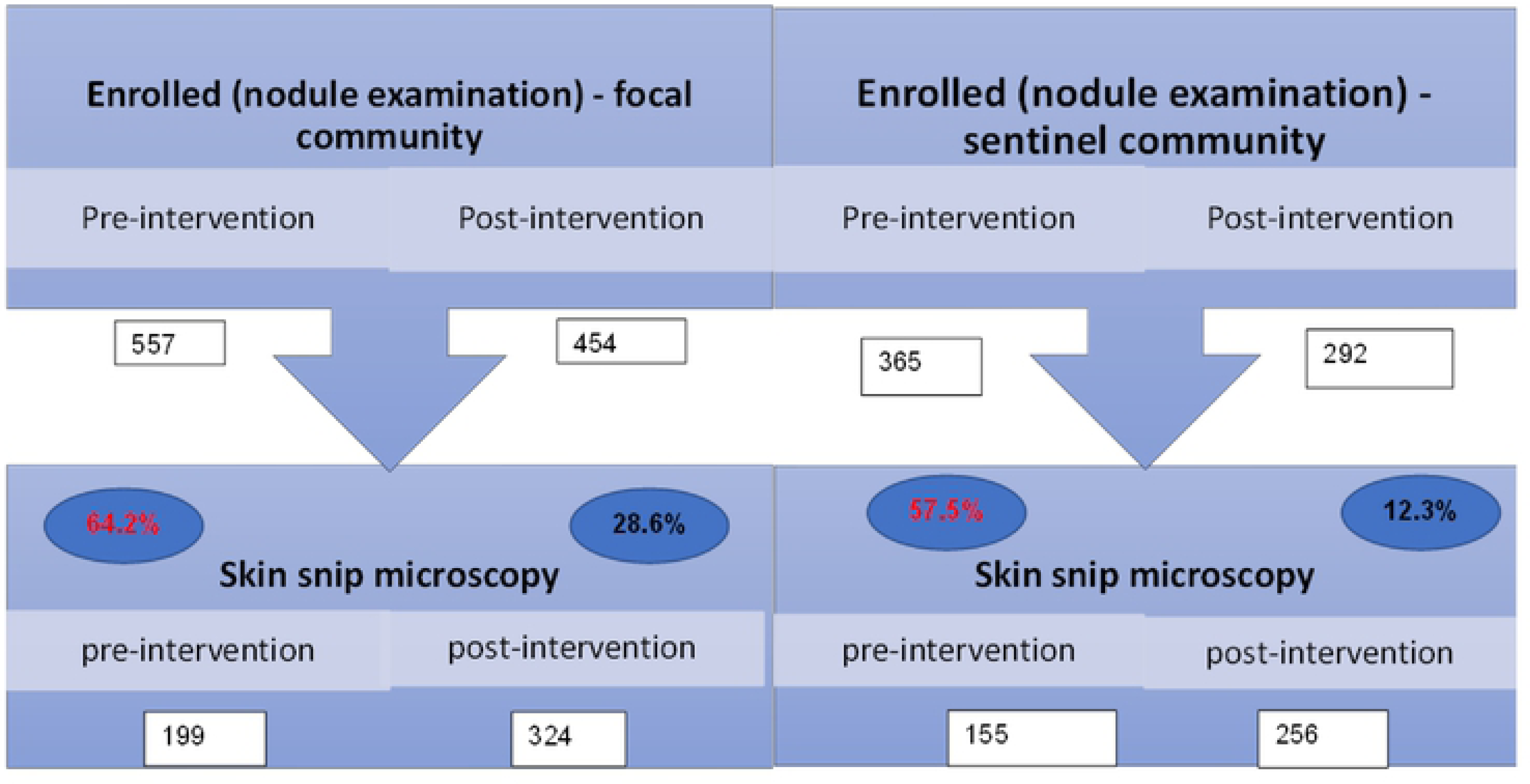
Chart showing population enrolled and screened before and after intervention.

A comparison of mf prevalence before and after the intervention in the focal communities showed a significant reduction (χ2: 40.1; p<0.001) from 35.7% (95%CI: 29.0 - 42.8) to 12.3% (95%CI: 9.0 - 16.4). In the sentinel community, the mf prevalence decreased by six percentage points from 11.0% (95%CI: 16.5-17.0%) to 5.1% (95%CI: 2.7-8.5%) (χ2=4.95; p=0.03).

Community and age were included in the final logistic regression model which shows the likelihood of having mf infection is 77% lower after the intervention compared to before (OR=0.22, 95%CI: 0.13 - 0.41, p=0.001), controlling for community and age (table 5).

CMFL was 0.66 mf/ss overall in the three focal communities and 0.13 mf/ss in the sentinel community before the AIS implementation. This was reduced to 0.27mf/ss and 0.11mf/ss respectively. The highest CMFL drop was recorded in Mankakoun - which decreased from 0.89 mf/ss to 0.49 mf/ss (table 7). The average mf intensity in the focal communities was 3.1 mf/ss before the intervention and 2.3 mf/ss after the intervention. In the sentinel community, the average mf intensity before intervention of 1.3 mf/ss significantly reduced to 0.7 mf/ss (p=0.03).

The mf intensity distributions in the plots (Fig 4) demonstrate the decrease in intensity distribution after intervention in the three focal communities, aside from 2 outliers (243mf/ss and 187mf/ss). Removing the two outliers from the sample resulted in a drop of post-intervention CMFL to 1.02 mf/ss and average mf intensity to 0.26.

**Fig 4.**
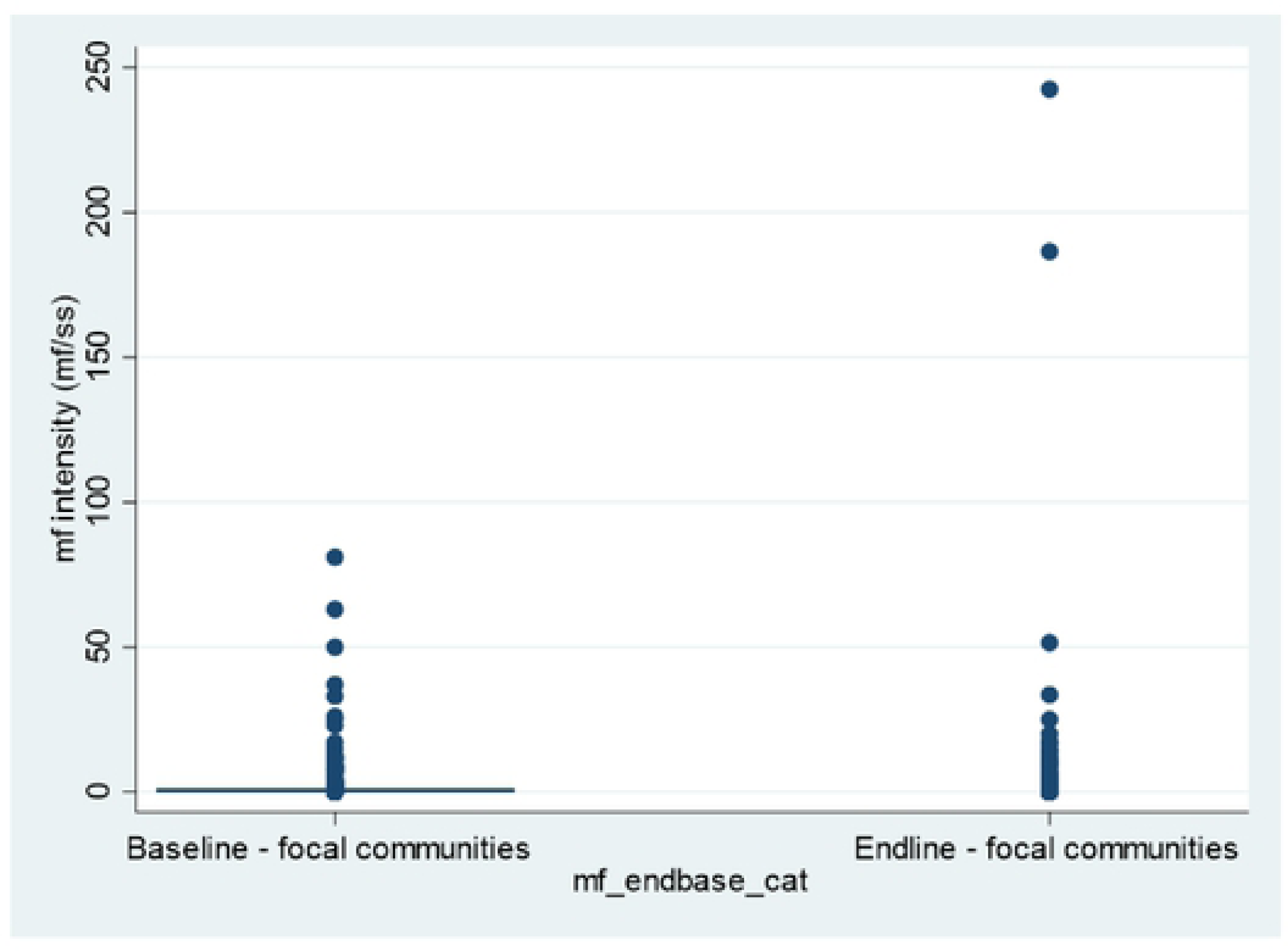
Mf intensity distribution in focal communities.

### Entomology

During the post-intervention transmission assessment, 20,792 flies were collected and 8,908 (43.0%) were dissected. Most of the flies caught were from Camp sic (39.0%; 8,106). A total of 454 flies were parous giving a parity rate of 5.1% of flies dissected for all the sites and months combined. This ranged from 3.9% in Mankare to 6.3% in the Makoupsap catch point, and September having the highest overall parous rate (all sites combined) (table 8). A total of 15 (3.3%) flies were infected among the 454 flies which were parous, with both Camp sic and Makouopsap having similar infection rates of 4.5%. The highest infection rate (14.3%) was observed in October at Mankare catching point. Ten (10) flies were found to be infective giving an infective rate of 2.2% of parous flies dissected.

**Table 8:** Entomological indices.

| Site | Months | # flies caught | # dissected | % of dissected flies parous (# parous) | % of parous flies infected (# infected) | % of parous flies infective (# infective) | MBR | MTP |
| --- | --- | --- | --- | --- | --- | --- | --- | --- |
| **Mankare (riverbank)** | August | 1,670 | 610 | 9.0% (55) | 0.0% (0) | 0.0% (0) | 12,942.5 | 0 |
|  | September | 3,076 | 764 | 2.1% (16) | 0.0% (0) | 0.0% (0) | 11,535.0 | 0 |
|  | October | 1,151 | 616 | 1.1% (7) | 14.3% (1) | 14.3% (1) | 8,920.3 | 14.5 |
|  | November | 1,709 | 1,429 | 3.8% (55) | 7.3% (4) | 7.3% (4) | 6,408.8 | 17.9 |
|  | **Total/Average** | **7,606** | **3,419** | **3.9% (133)** | **3.8% (5)** | **3.8% (5)** | **\* 9,951.6** | **\*8.1** |
| **Camp sic (riverbank)** | August | 1,886 | 599 | 11.4% (68) | 8.8% (6) | 4.4% (3) | 14,616.5 | 73.2 |
|  | September | 3,215 | 517 | 10.3% (53) | 0.0% (0) | 0.0% (0) | 12,056.3 | 0 |
|  | October | 1,082 | 464 | 3.4% (16) | 0.0% (0) | 0.0% (0) | 8,385.5 | 0 |
|  | November | 1,923 | 1,209 | 1.6% (19) | 5.3% (1) | 5.3% (1) | 7,211.3 | 6.0 |
|  | **Total/Average** | **8,106** | **2,789** | **5.6% (156)** | **4.5% (7)** | **2.6% (4)** | **\*10,567.4** | **\*19.8** |
| **Mankakoun (community)** | August | 0 | 0 | 0.0% (0) | 0.0% (0) | 0.0% (0) | - | 0 |
|  | September | 2,145 | 499 | 6.6% (33) | 0.0% (0) | 0.0% (0) | 8,043.8 | 0 |
|  | October | 790 | 502 | 6.6% (33) | 0.0% (0) | 0.0% (0) | 6,122.5 | 0 |
|  | November | 1,203 | 1,006 | 5.5% (55) | 1.8% (1) | 1.8% (1) | 4,511.3 | 4.5 |
|  | **Total/Average** | **4,138** | **2,007** | **6.0% (121)** | **0.8% (1)** | **0.8% (1)** | **\*4,669.4** | **\* 1.1** |
| **Makouopsap**<br>**(community)** | August | 73 | 51 | 17.6% (9) | 11.1% (1) | 0.0% (0) | 565.8 | 0 |
|  | September | 35 | 8 | 62.5% (5) | 0.0% (0) | 0.0% (0) | 131.3 | 0 |
|  | October | 89 | 43 | 14.0% (43) | 0.0% (0) | 0.0% (0) | 689.8 | 0 |
|  | November | 745 | 591 | 4.1% (24) | 4.2% (1) | 0.0% (0) | 2,793.8 | 0 |
|  | **Total/Average** | **942** | **693** | **6.3% (44)** | **4.5% (2)** | **0.0% (0)** | **\*1,045.1** | **\*0** |
|  | **Overall**<br>**Total/Average** | **20,792** | **8,908** | **5.1% (454)** | **3.3% (15)** | **2.2% (10)** | **\*\*6,558** | **\*\*7.3** |

*\*Average of months per site*
*\*\*Overall average for all sites and months*

The average MBR for all sites and months was 6,558 bites/man/month: Camp sic site having the highest. In both Camp sic and Mankere catching points, August recorded the highest MBR compared to the other months. In Makouopsap, the MBR was much higher in November. The average MTP for all sites and months was 7.3 infective larvae/man/month, highest at Camp sic (19.8 infective larvae/man/month). August in Camp sic observed the highest MTP of 73.2 infective larvae/man/month).

## Discussion

The aim of this study was to evaluate the impact of AIS package implemented over a period of two years to accelerate the progress towards onchocerciasis elimination in the Massangam HD. This is an area noted for high onchocerciasis prevalence and ongoing transmission after more than 20 years of annual IVM (15, 17). Parasitologically, we compared the mf prevalence, nodule prevalence, CMFL and mf intensity between a pre- and post-intervention surveys. Parity, infection, infectivity, MBR and MTP were determined to ascertain whether transmission was still ongoing. Interventions were also compared to address challenges for better implementation of subsequent rounds.

The AIS resulted in accelerated progress towards elimination as compared to annual CDTi over longer time period, with a reduction of prevalence of 29.0% under AIS in 2017-2019 compared to 14.6% with IVM in 2011-2015. Similarly, in Massangam, there was a reduction of mf prevalence by 23.2% following the two years of AIS compared to 20.3% reduction over 15 years of treatment with IVM (1996-2011) (15). Similar results in accelerating elimination using doxycycline treatment as an “end-game strategy” were reported in Brazil and Venezuela (23). Doxycycline has curative and macrofilaricidal effects through targeting symbiotic *Wolbachia* (20, 24-26), compared to IVM which is only microfilaricidal, targeting the mf.

Implementation of TTd was feasible with good adherence to treatment for both interventions as seen by treatment completion rates of more than 90%. Similar treatment compliance rate was reported by Wanji and colleagues who indicated 97.5% of the people who started treatment completed it (25). Apart from doxycycline being a broad-spectrum antibiotic that could have had additional benefits on participants, distribution of the drug was accompanied with meal support given to the participants during the treatment course and using support from CDDs through DOT which enhanced adherence. The major challenge was the poor uptake in testing – primarily due to issues over fear and pain associated with the skin biopsy test used. There is need to find an alternative test which is less painful and ideally more sensitive, or alternatively use anybody test in treatment of naïve areas. Another factor for the low screening rate was fatigue over requests for multiple rounds of testing from baseline through the interventions and impact assessment. However, offering two rounds of TTd was important, in particular in this setting with mobile populations and uncertain denominators, which meant that additional rounds ensured new people moving to the area or who missed prior rounds were captured. It also meant those initially fearing to take the test had an opportunity to see the benefits of the new approach from those involved in the first round. Having highly mobile populations also means the time from census to test to treatment should be kept as short as possible.

Larviciding was also successfully implemented with a reduction in fly populations seen through the early and later weeks of larviciding period. However, during both rounds of larviciding, the rise in fly population observed around the fourth week could be explained by influx of blackflies from other breeding sites which were not being treated (as they were not productive by the time of prospection or due to complexities in the river system). Following change of treatment spots upstream to cover these new breeding sites, real-time monitoring, recalculation of larvicide quantities, the fly population experienced. This is typical of the challenges in complex breeding sites/river systems as in the case of River Mbam. Thus, it is important to monitor fly populations and larvae to allow for adaptations of temephos concentrations as needed.

Biannual MDA was also successfully implemented but the gains here were more associated to the added opportunity of eligible populations to receive at least one round of IVM (improving overall % that received at least one round) to impact on perennial transmission rather than perhaps about ensuring everyone received two rounds in a year - which was really the primary purpose. However, each round recording a higher coverage from the previous one could be an indication of increased reach. The difference in the reported and program treatment coverages could be suggestive of National Program sometimes over-reporting coverages during MDAs, thus the need for CES to validate program coverage. Kamga et al (2018), reported a similar observation where the survey coverages in three HDs were lower than the program coverages with a significant difference varying from 14.1% to 22.0% (27).

Although nodule prevalence was conducted during the post-intervention survey, this was used only to compare with the before prevalence, as nodule palpation was useful in mapping the distribution of onchocerciasis to identify priority areas for OV infection prevention activities (28) and not as a tool to evaluate progress towards elimination (29). This however after two years of implementation decreased like all other indicators in both the three focal and sentinel communities which could also be suggestive of the effect of doxycycline (macrofilacidal) on adult worms as they usually live in subcutaneous nodules and produce mf, which are found throughout the individual’s body (30). However, this finding is not conclusive, as usually time is needed for the nodules to disappear after the death of the adult worm. In the sentinel community, an additional explanation could be the fact that the treatment with IVM was conducted biannually, which is more effective than the annual treatment in preventing the appearance of nodules (31). It is also important to note that not only are nodules not a good measure to access elimination progress, but they are also difficult to find (particularly in women) and they can be indicative of other diseases.

Entomological assessment demonstrates that transmission is still ongoing in this area (1.1 infective flies per 1000). This can be expected based on the timeline of the interventions and expected time to impact. It serves to highlight the challenges in interrupting OV transmission in an environment with complex breeding sites and open transmission zones. It is very likely that there was repopulation of flies from the neighbouring Centre region, which has very high on-going transmission and was not targeted in the study. Thus, optimal timing of impact of larviciding with treatment to impact on transmission needs to be further explored. Furthermore, pool screening was not used to estimate the infective rate (as per WHO guideline), so likely underestimated the true rate of infection. However, the low parous rates in the study area are suggestive of a young population of flies, and this is likely related to recolonisation of the sites following larviciding activities. Furthermore, larviciding was done during the dry season where fly population and activity are usually lower compared to rainy seasons noted as peak transmission seasons, which further lowered the population.

### Limitations

There are a number of limitations that need to be taken into consideration when interpreting the results of this study.

Firstly, the study design does not have a comparator. We could not compare the results with another similar area where only IVM was distributed annually, as such an area with similar transmission dynamics does not exist. Also, it is difficult to disentangle the impact of each approach whether TTd, biannual IVM or vector control. However, the study saw a bigger decrease in two years than over a longer period of treatment with annual CDTI, as also demonstrated by a similar study conducted in the Southwest Region of Cameroon, having a great impact (32).

Secondly, sample size issues might have affected prevalence estimates. Although all eligible individuals were invited to participate, the sample size was affected by a lot of resistance to testing children as well as general acceptability issues related to the multiple screenings in this area, with the endline representing the fourth time skin snips were being taken in the communities in 2 years. To monitor trends outside the focal area, only one sentinel site was used due to budget limitations. A wider survey of a selection of the surrounding communities would also have been useful. During the time of the study, it was clear that the semi-nomadic populations in the communities were not adequately being reached by the programme. As such, a separate test and treat intervention targeting this hard-to-reach group was designed and implemented. The results of this are not included in this paper and the population and their role in potential facilitation of on-going transmission may be under-represented in this study.

The integration with the ongoing IVM programme, the test and treat nor the endline survey could not be optimally timed. In particular, implementing an endline survey after more than a year (33) could have allowed for more time for doxycycline effect after the second round of test and treat. Mobility of the population also made it very difficult to get good population estimates and with new people moving into the area would have potentially brought infection in and confounded results.

Finally, the indices of transmission at baseline used a slightly different methodology and were conducted at different seasons meaning it is not useful to directly compare with the endline assessments. However, the entomological indices were still useful in understanding if transmission is on-going. Future timing and methodologies for entomological assessments must be in line with the endline.

## Conclusion

To conclude, the findings support the WHO recommendation of using alternative strategies to facilitate progress towards elimination of onchocerciasis as seen in the case of the Massangam HD. Despite the challenges in implementation, especially coverage, the impact of these strategies was still significant with the reductions in mf prevalence, CMFL as well as mf intensity in both focal communities and sentinel community. We therefore recommend its use in similar transmission zones with persistent high prevalence. The use of test and treat with doxycycline has a particular benefit in that it is not contraindicated in *Loa loa* infected persons, unlike for IVM and as it is potentially curative, requires less rounds of administration to impact on *O. volvulus* transmission and therefore can be beneficial in hard-to-reach communities where it is difficult to consistently reach with high coverage of IVM. There is need to consider the above limitations and implementation challenges (34), as well as the problem of how to sustain the impact of AIS in a complex transmission zone with neighbouring areas of high transmission, needs to be explored. The potential to conduct a similar intervention in the Centre region could be worthwhile.

## Data Availability

All relevant data are within the manuscript.

## Acknowledgements

We thank the National NTDs department in Cameroon, the West Regional Delegation of Public Health, and the Massangam Health District for facilitating the implementation of this research. We also thank the communities for accepting to take part in this work. Thank you goes to our partner The Filariasis and other Tropical Diseases Research Center (CRFilMT) for their technical and laboratory support. We equally thank Alex Chailloux for producing the map showing the various communities, and Susan D’Souza and Richard Selby for reviewing the manuscript.

